# Use of mobile health apps in low-income populations: a prospective study of facilitators and barriers

**DOI:** 10.1101/2019.12.22.19015636

**Authors:** Patrick Liu, Katia Astudillo, Damaris Velez, Lauren Kelley, Darcey Cobbs-Lomax, Erica S. Spatz

## Abstract

**Background:** Mobile applications (apps) are increasingly popular in healthcare. For low-income populations, barriers exist, yet limited data are available about the challenges and catalysts for adoption.

**Methods and Results:** We partnered with a primary care center and a community organization and recruited patients to use a health app. A community health worker (CHW) consented participants, downloaded the app and instructed on its use, and provided ongoing technical support. Bi-weekly surveys for three months were sent via email/text to assess participant experiences and perceptions.

The majority (81 of 108 [75.0%] English language-preferred and 50 of 52 [96.2%] Spanish language-preferred) of patients approached were enrolled. Common reasons for declining were: did not own a smartphone (13.8%), did not have email (20.7%), and not interested (58.6%). Enrollment challenges included: insufficient storage, unfamiliarity with downloading apps, forgotten passwords to email accounts, and slow/absent WiFi connection – which the CHW and the app company were able to address. Most participants, English and Spanish language-preferred respectively, were interested in monitoring their health through an app (74.4%; 70.4%), connecting devices such as FitBits© and blood pressure cuffs (78.9%; 50.0%), and being the owner of their health records (83.6%; 95.6%). There were concerns about sharing health information with research teams (66.7%; 51.9%), and data being sold (83.0%; 70.4%). However, many (58.6%; 87.2%) reported being likely to share health data with a trusted research team. Compared with before the study, most felt more comfortable using health apps (67.4%; 82.1%) and more likely to participate in research using apps (76.2%; 72.4%).

**Conclusions:** The assistance of a CHW facilitated the enrollment of low-income individuals in a mobile health app by fostering trust and sustained engagement. Participants were interested in having several app features. Despite concerns about data privacy, they demonstrated greater interest in mobile health app use and research participation at study conclusion.

## INTRODUCTION

Applications (apps) accessible on digital platforms are increasingly popular in healthcare settings, especially with mobile device ownership surpassing 90% and smartphone penetrance estimated at approximately 60% in the United States in 2013 (1, 2). Apps have the potential to help patients engage more with their health through the monitoring of health behaviors (e.g., step counts; physical activity; dietary habits) and physiologic parameters (e.g., heart rate; blood pressure; blood glucose level) by connecting external devices. Some health apps provide patients access to their medical data through patient portals and opportunities to participate in research using their smart devices. Other mobile health (mHealth) apps aim to support adherence to treatments and help providers encourage treatment between healthcare visits, though literature has not shown consistently significant impacts (3, 4). For low-income populations, however, unique barriers to mHealth utilization exist. These barriers include fluency with mobile apps, limited health literacy, lack of empowerment, and historical mistrust of healthcare systems (5-8). As mHealth platforms play a larger role in healthcare delivery, the digital divide could serve to worsen health disparities (9-11).

mHealth apps that are sensitive to the user needs of vulnerable populations have the potential to gain uptake in more diverse communities. Specifically, the visual and linguistic design of mHealth apps, along with considerations for *how* mHealth apps are introduced to patients – may increase adoption and acceptability (12, 13). Recent literature on mHealth interventions has primarily focused on mHealth adoption among health workers to facilitate their work and interactions with patients, primarily in low- and middle-income countries (14-16). Some of these studies use community health workers (CHW) as frontline providers to connect with patients and improve their health outcomes (17-19). Ideally, because of their shared lived experiences and unique positions within the healthcare team, CHWs have the opportunity to improve integration of mobile health apps into the health and healthcare experience of their patients. However, little literature exists regarding digital health uptake among low-income populations in the United States, and how CHWs may facilitate that process. Furthermore, little is known about this population’s attitudes towards mobile health apps, the ability of apps to increase access to and sense of ownership of medical information, and how patients’ perceptions may change over time with consistent use of an mHealth app with technological support from CHWs.

To understand the real-world challenges and catalysts for adoption of health-related mobile technologies among low-income patients in the United States, we evaluated the experiences of underserved patients engaged with an mHealth app. We aimed to learn how a digital health platform should be configured to be valuable for underserved patients and integrated into healthcare settings to facilitate mHealth adoption with sustained engagement.

## METHODS

We conducted a community-based participatory research project in partnership with a community health organization, a primary care center, and a digital health company. Project Access-New Haven (PA-NH) is a community health organization that provides intensive patient navigation to improve access to care for underserved individuals in the Greater New Haven area. The Yale Primary Care Center (PCC) is a hospital-based clinic serving primarily patients with Medicaid; the Center offers a variety of integrated services, including general primary care, nutrition, addiction, behavioral health, home visitation, and refugee health. Hugo Health is a digital health company that produced its eponymic app, which provides its users access to medical records from different healthcare systems, allows integration of wearable and other cloud-based devices (e.g., Fitbits, wireless scales, and wireless blood pressure cuffs), and facilitates participation in healthcare research. With patients’ permission, patients can opt to share portions of their data with research teams and answer surveys from researchers using their email and/or smart devices.

### Study Design and Implementation

Our study’s first phase involved designing an implementation plan that optimized the capabilities of PA-NH community health workers and Hugo with the following goals: (1) to create a seamless experience with study enrollment; (2) to refine the research aims and identify key areas of interest to be assessed in the questionnaires; (3) to collaboratively develop and cognitively test survey questions in both English and Spanish to improve the quality of the questions and their comprehension. We held regular phone meetings for all stakeholders (PA-NH, Hugo Health, Yale PCC, and Yale researchers). The second phase included patient enrollment and study participation. We obtained Institutional Review Board approval of the protocol (ID 2000022503), and all patients provided informed consent.

### Participant Recruitment, Eligibility, and Enrollment

We recruited patients from PA-NH and the Yale PCC. We aimed to enroll 80 English language-preferred patients and 50 Spanish language-preferred patients to participate in the study for three months. Patients attending primary care visits were eligible if they had ongoing healthcare needs for which the Hugo app would be helpful, demonstrated basic literacy in English or Spanish, and had access to a smart device. A CHW stationed at the healthcare centers assessed eligibility, gauged interest in study participation, and consented participants for enrollment. If criteria were met but patients did not want to participate in the study, the CHW logged the reason(s) for non-participation (i.e., lack of smart phone, lack of email or access to email, lack of interest, or other). If inclusion criteria were met and patients consented to participating in the study, the CHW then downloaded the app on the patients’ mobile devices, connected them to their health portal(s), and instructed them in the use of different features of the app that may have been the most applicable to the patient (e.g., following up on test results, sharing health data with clinicians/family members; tracking symptoms and physiologic data from connected devices). The CHW then practiced using these different functionalities with the patient. Finally, the CHW kept a qualitative log of barriers encountered and strategies utilized to address those barriers during the enrollment process. These logs were discussed on weekly calls with all stakeholders.

### Survey Development and Dissemination

A baseline survey was administered at enrollment to gather participants’ background characteristics, followed by six brief surveys administered through the Hugo app on a bi-weekly basis. The first five follow-up surveys assessed participants’ interest and concerns about using health apps, data access and sharing, and participation in research. The sixth and final survey assessed patients’ experience with the health app and interacting with the CHW, as well as how, if at all, their knowledge and attitudes about health apps changed through this experience. Links to the six surveys were sent via email or text according to patient preference. Responses were archived in Hugo.

The CHW provided ongoing tech support, with the help of Hugo staff, throughout the study by phone call, text, and in person. The CHW also sent reminders to complete surveys at one and three months after enrollment. Monetary incentives were provided at enrollment and three months, after patients completed their baseline and final surveys. Incentives were not offered for responding to the first five follow-up surveys.

### Feedback After Completion of Study

We sought feedback from 15 English language-preferred and five Spanish language-preferred participants at the end of their enrollment in the study. Study participants were randomly selected. We asked them questions regarding (1) their experiences with their participation in the study, including what they did and did not like; (2) what was difficult using Hugo and what could have made the experience better; (3) whether and how having a community health worker was helpful; (4) if they feel differently about owning/having access to their medical information; (5) whether they feel differently about using an app to monitor health; and (6) if they feel differently about participating in research using an app.

### Main Outcome Measures

Our main outcome measures were related to: (1) patients’ willingness and comfort with using mobile apps for a) health monitoring, b) healthcare, and c) research participation; (2) facilitators and barriers to utilizing an mHealth app; (3) utility of a CHW to facilitate mHealth engagement; and (4) principal uses and functionality of interest to participants.

### Statistical Analysis

We used descriptive analyses to report responses to our surveys. All analyses were stratified by preferred language (English vs. Spanish). Excel, version 14.7.6 (Microsoft Corp.) and RStudio, version 1.1.423 (RStudio Inc.) were used for all analyses.

## RESULTS

### Patient Enrollment

Among 108 English language-preferred patients and 52 Spanish language-preferred patients approached for the study, 81 (75.0% of 108) and 50 (96.2% of 52) were enrolled, respectively (**Table 1**). The mean income of English language-preferred patients was 113% of federal poverty level (FPL), and were primarily Black/African American (74%), followed by Hispanic/Latino (16%), White (7%), and Other (2%). Spanish language-preferred participants had a mean income of 82% FPL and all (100%) were Hispanic/Latino. Approximately one-third of both English. (34.6%) and Spanish (34.0%) language-preferred respondents were assessed as having low health literacy.

**Table 1:** Demographic characteristics of English-preferred and Spanish-preferred patients enrolled in the study. <u>Footnotes:</u> * Assessed in-person using the Short Assessment of Health Literacy in English and Spanish

|  | English-preferred (n=81) | Spanish-preferred (n=50) |
| --- | --- | --- |
|  | No. (%) | No. (%) |
| **Gender** |  |  |
| Female | 59 (72.8%) | 32 (64.0%) |
| Male | 22 (27.2%) | 18 (36.0%) |
| **Age** |  |  |
| Years (mean) | 48.2 | 46.4 |
| **Race/Ethnicity** |  |  |
| Black/African American | 61 (74.4%) | 0 (0.0%) |
| Hispanic/Latino | 13 (15.9%) | 50 (100.0%) |
| White | 6 (7.3%) | 0 (0.0%) |
| Other | 2 (2.4%) | 0 (0.0%) |
| **Household Income** |  |  |
| % FPL (mean) | 113 | 82 |
| **Education** |  |  |
| Less than elementary | 1 (1.2%) | 7 (14.0%) |
| Elementary | 2 (2.4%) | 12 (24.0%) |
| Some high school | 6 (7.3%) | 6 (12.0%) |
| High school/GED | 47 (57.3%) | 17 (34.0%) |
| Some college | 22 (26.8%) | 5 (10.0%) |
| Associate degree | 2 (2.4%) | 1 (2.0%) |
| Bachelor's degree | 2 (2.4%) | 2 (4.0%) |
| **Employment** |  |  |
| Employed full-time | 19 (23.2%) | 8 (16.0%) |
| Employed part-time | 18 (22.0%) | 20 (40.0%) |
| Unemployed - looking | 14 (17.1%) | 10 (20.0%) |
| Unemployed - not looking | 6 (7.3%) | 9 (18.0%) |
| Unable to work | 25 (30.5%) | 3 (6.0%) |
| **Health Literacy\*** |  |  |
| Low ($\leq 14$) | 28 (34.6%) | 17 (34.0%) |
| Adequate ($> 14$) | 53 (65.4%) | 33 (66.0%) |

The most common reasons cited by the 29 patients who declined participation were: did not have access to a smartphone (4 of 29), did not have email (6 of 29), and not interested (17 of 29). Technological challenges to enrollment of study participants included: insufficient storage; unfamiliarity with downloading and using apps; forgotten passwords to email accounts; infrequent email use (resulting in a text option for communication); and slow/absent WiFi connection. Enrollment of English language-preferred patients began on April 25, 2018 and ended May 31, 2018, and enrollment of Spanish language-preferred patients started on October 11, 2018 and ended December 20, 2018.

### Survey Response Rates

Survey response rates ranged from 48.2% (2-week survey) to 86.4% (12-week survey) of 81 English language-preferred participants, and 50.0% (2-week and 8-week survey) to 94.0% (12-week survey) of 50 Spanish language-preferred participants (**Appendix Table 1**). The highest response rates were for the surveys sent at one month and three months after enrollment. *Patients’ Willingness and Comfort with Using the Features of Mobile Apps*

On the second follow-up survey, sent four weeks into the study, there were 78.9% (41 of 52) and 50.0% (16 of 32) of English language-preferred and Spanish language-preferred respondents, respectively, who felt comfortable sharing the information gathered on connected devices, such as FitBits**^©^** and blood pressure cuffs, to a trusted research team (**Figure 1**).

**Figure 1:**
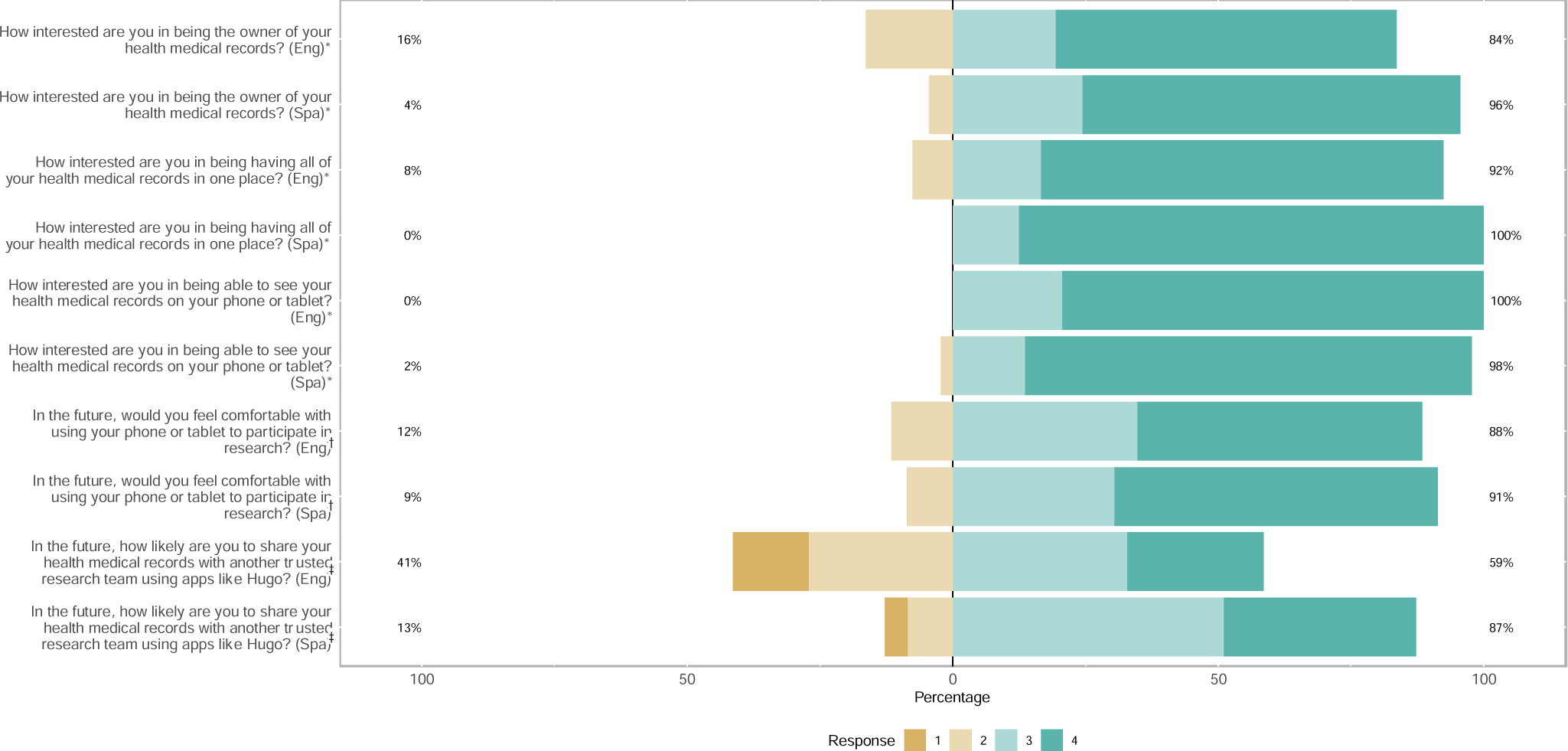
Baseline interest and comfort with using the features of mobile apps, by preferred language. <u>Footnotes:</u> * Survey options: “Very interested,” “Somewhat interested,” “Not sure, I would need more information before I could answer,” “Not at all interested” † Survey options: “Very likely,” “Somewhat likely,” “Not sure, I would need more information before I could answer,” “Not at all likely” ‡ Survey options: “Very comfortable,” “Somewhat comfortable,” “Not sure, I would need more information before I could answer,” “Not at all comfortable”

On the final survey, conducted three months after enrollment, the vast majority were very or somewhat interested in being the owner of their health records (83.6% of 67; 95.6% of 45), having their health records in one place (92.4% of 66; 100.0% of 40), and being able to view them on their phones or tablets (100.0% of 68; 97.7% of 44) (**Figure 1**). Furthermore, 88.4% (61 of 69) English language-preferred participants and 91.3% (42 of 46) Spanish language-preferred participants indicated that they felt somewhat or very comfortable with participating in research using their mobile phone or tablets.

Additionally, at the end of the study, 58.6% (41 of 70) and 87.2% (41 of 47) reported being somewhat or very likely to share health data with a trusted research team (**Figure 1**). Compared with before the study, 67.4% (29 of 43) and 82.1% (23 of 28) reported feeling more comfortable using health apps, and 76.2% (32 of 42) and 72.4% (21 of 29) reported being more likely to participate in research using apps (**Figure 2**). Moreover, 35.7% (25 of 70) and 17.8% (8 of 45) respondents stated that they used other mobile health apps during their enrollment in the study (**Appendix Table 2**).

**Figure 2:**
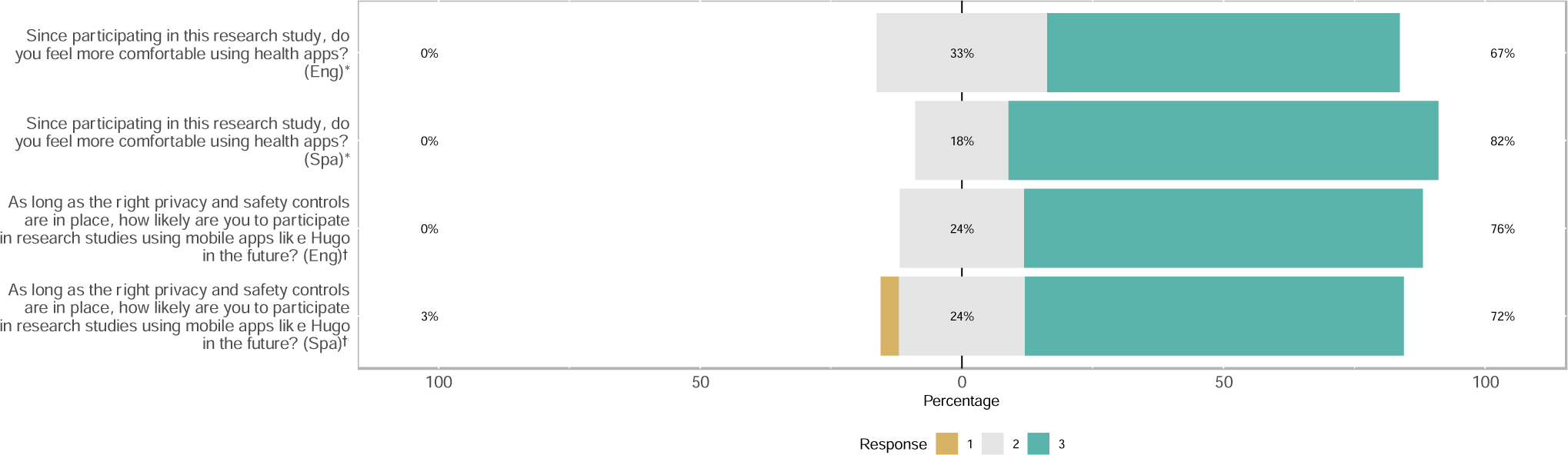
Post-participation interest in health apps, by preferred language. <u>Footnotes:</u> * Survey options: “I am now more comfortable using health apps,” “I have the same level of comfort using health apps (no change in comfort),” “I am now less comfortable using health apps” † Survey options: “More likely than before this research study,” “Same likelihood as before this research study,” “Less likely than before this research study”

### Facilitators and Barriers to Utilizing an mHealth App

Most participants indicated that it was preferable to answer survey questions when they are texted to them (55.8% of 43; 75.9% of 29) rather than emailed to them (34.9% of 43; 10.3% of 29) (**Appendix Table 2**). Additionally, as further detailed below, most participants felt the support of a CHW was helpful for understanding and using the mHealth app.

Among the 14 English and 12 Spanish language-preferred respondents who stated that they did not use Hugo by the end of the study, the most common reasons cited were did not have a reason to log on (35.7%; 25.0%); could not figure out how to use the app (14.3%; 50.0%); forgot about the app (35.7%; 16.7%); could not remember their username or password (14.3%; 25.0%) (**Appendix Table 2**).

Regarding privacy policies, the majority of participants (82.6% of 46 English language-preferred; 92.6% of Spanish language-preferred respondents) either strongly agreed or agreed with being interested in learning more about privacy policies that explain how data are collected and used via digital health platforms to participate in research (**Figure 3**). Fewer respondents (50.0% of 46; 46% of 25) indicated that, if they trust the research team, they do not need to read or hear about privacy policies when using digital health platforms to participate in research. Only a minority (33.3% of 45; 19.1% of 21) strongly agreed or agreed that privacy policies should be shorter, even if some details about the policy are left out. Most (80.0% of 45) English language-preferred respondents, but (21.4% of 14) few Spanish language-preferred respondents strongly disagreed or disagreed with the statement, “I will never read privacy policies even if they were made shorter and easier to understand.”

**Figure 3:**
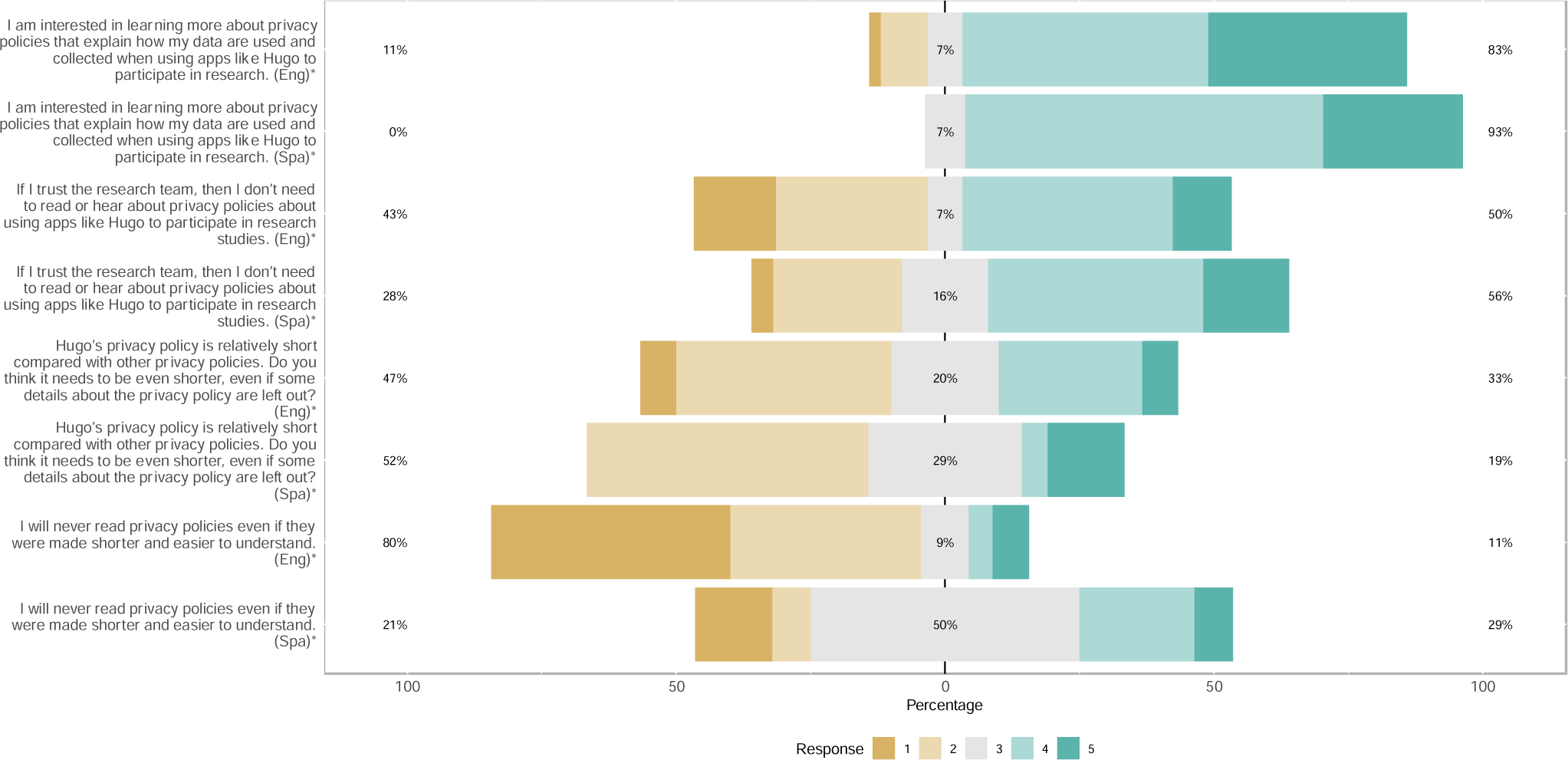
Privacy policy concerns, by preferred language. <u>Footnotes:</u> * Survey options: “Strongly agree,” “Agree,” “Not sure,” “Disagree,” “Strongly disagree”

Most participants were somewhat or very concerned about sharing health information with research teams (66.7% of 48 English language-preferred; 51.9% of 27 Spanish language-preferred respondents), and most had concerns that information collected through apps could be sold or shared without their permission (85.9% of 47; 70.4% of 27) (**Figure 4**).

**Figure 4:**
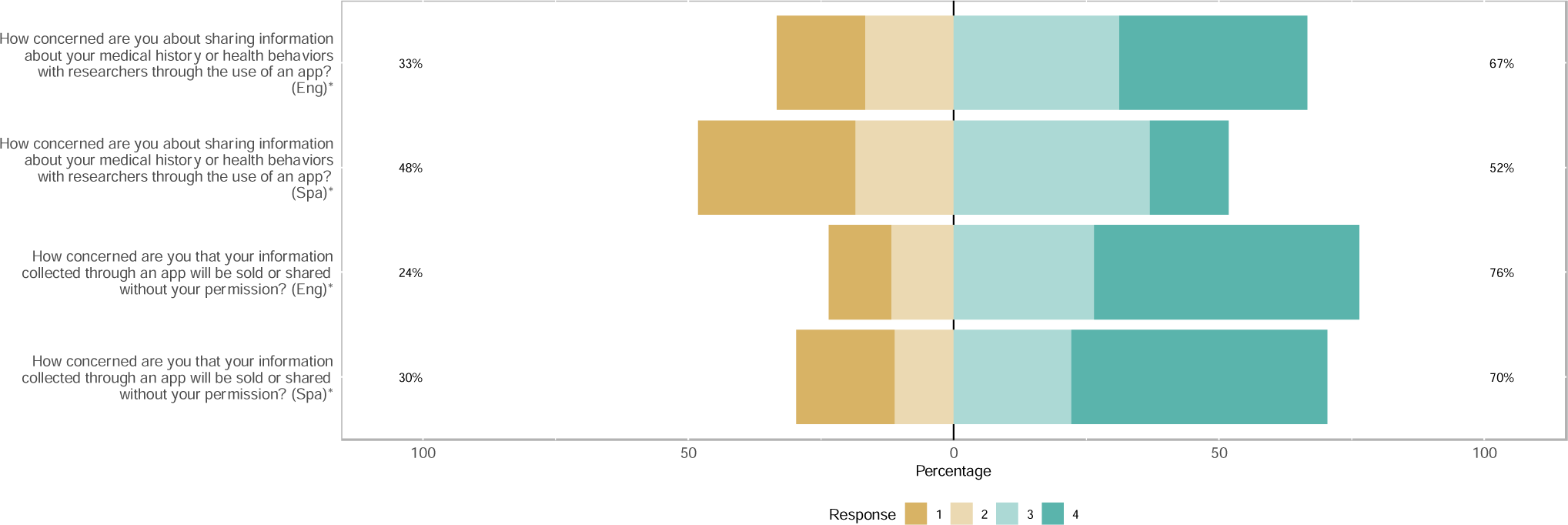
Data sharing concerns, by preferred language. <u>Footnotes:</u> * Survey options: “Very concerned,” “Somewhat concerned,” “Not sure,” “Not at all concerned”

### Utility of a CHW to Facilitate mHealth Engagement

Requests to speak with the CHW about any questions regarding features of the digital health app were consistently higher for Spanish language-preferred participants than for English language-preferred participants (across all six survey periods, the median percentage of patients requesting assistance was 51.6% for Spanish language-preferred participants and 28.1% for English language-preferred respondents) (**Figure 5**). Up to 48.8% of English language-preferred (10-week survey) and 63.0% of Spanish language-preferred respondents (6-week survey) indicated a desire to speak with the CHW about features of the digital health app. Furthermore, 60.9% of 46 English language-preferred respondents and 79.2% of 24 Spanish language-preferred respondents strongly agreed or agreed with being interested in having someone from PA-NH or the research team explain the app’s privacy policies to them again or in greater detail.

**Figure 5:**
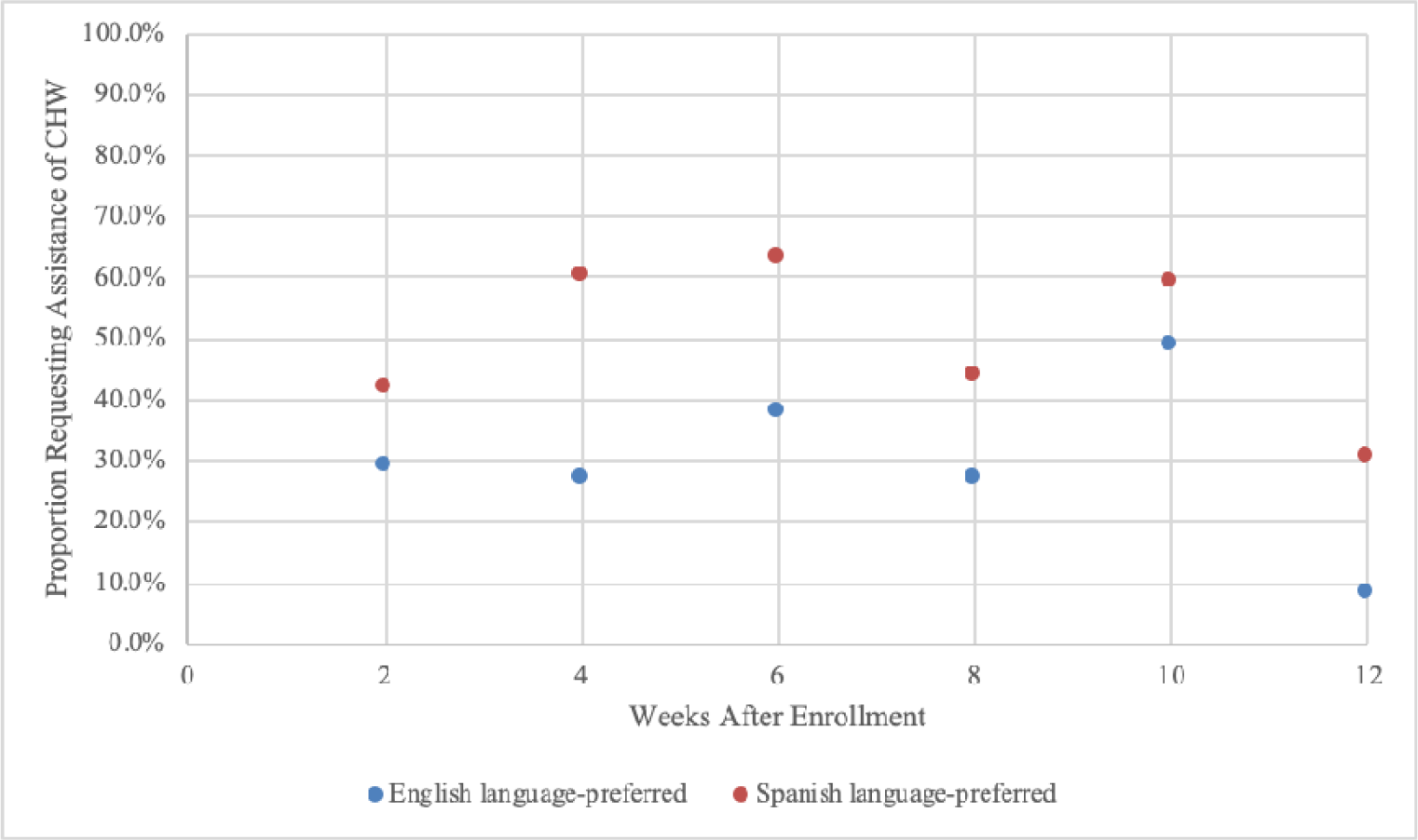
Proportion of participants requesting the assistance of a community health worker on each follow-up survey, by preferred language.

### Principal Uses and Functionalities of Interest to Participants

At the beginning of the study, English language-preferred respondents (86.5% of 53) and Spanish language-preferred respondents (86.7% of 26) found the digital health platform most useful for keeping track of their own health information (**Appendix Table 2**). There was specific interest in using the app to track their vital signs, symptoms post-surgery, and device (e.g., pacemaker) malfunctions. At the end of the study, participants used the app most often for connecting to medical records (48.6% of 70; 53.2% of 47); checking lab results (41.4% of 70; 44.7% of 47); and participating in research (35.7% of 70; 17.0% of 47).

Regarding what is most important to participants when deciding whether to participate in research, responses varied by cohort. Among English-preferred respondents, the most important attributes in descending order were: whether the research would benefit them or others; whether the research has any risk to them; getting paid for participation in the study; and having someone to contact if they have questions (**Appendix Table 2**). Among Spanish-preferred patients, the most important considerations for research participation in descending order were: whether the research would benefit them or others; having someone to contact if they have questions; and getting paid for participation in the study.

Furthermore, participants responded that they were interested in using apps that bring together people who share similar health conditions (65.1% of 43 English language-preferred; 74.1% of Spanish language-preferred patients), learning about other people’s experiences (71.4% of 28; 31.6% of 19, and learning about research studies and clinical trials (57.1% of 28; 10.5% of 19) (**Appendix Table 2**).

### Feedback After Completion of Study

Most surveyed individuals had a positive experience, reported that they trusted the community health worker and Hugo team, and were excited to use many of the features, including to view medical records and to participate in research. Select responses are displayed in **Appendix Text 1**.

## DISCUSSION

In our study of 81 English language-preferred and 50 Spanish language-preferred patients, we found marked interest from participants in using a digital health platform to monitor health (e.g., through aggregating health records into one location, checking lab results, and connecting external devices) and participate in research. Most, but not all, patients approached for the study owned a smart device, were excited about enrolling in the study, and responded to the surveys. There were concerns about the privacy of information gathered through digital health platforms, and a majority of participants were interested in having the CHW or another member of the research team review the privacy policies in greater detail. By the conclusion of the three-month enrollment, there was substantial interest among participants in being the owner of their health records, monitoring health through the app and connected cloud-based devices, and participating in research using digital health platforms in the future. A CHW was able to facilitate the efficient enrollment and teaching/utilization of the digital health platforms by addressing basic technical issues during the baseline visit and by elevating more complex questions and challenges to the digital health company, Hugo, and the research team at large.

Despite the proliferating ownership of cellular devices and development of digital health platforms in the United States, relatively little work has been done on identifying the needs of low-income populations to tailor mHealth interventions. Previous studies of vulnerable populations have found similar, if not increased, rates of mobile engagement through text messaging compared to the general population; therefore, text messaging may be an effective way of reaching out to this population (2, 20). A randomized controlled trial of 15 case managers and 67 patients in New York City leveraged a two-way text messaging system to demonstrate increased appointment and medication adherence (21). However, the use of mHealth apps has been criticized for not effectively taking into account the economic and social contexts of low-income populations. An mHealth app (myHealthButton) developed for Michigan’s Medicaid population was only actively used by approximately 1,500 of 2 million Michigan residents enrolled in Medicaid and Child Health Insurance Program (22). When Arizona’s governor announced that a mobile app would be developed for Arizona’s Medicaid beneficiaries, there was concern by Health Net, a plan overseeing 1.7 million Medicaid beneficiaries in Arizona and California, and others, that the app would not have a significant number of active users (22). When digital health platforms and mHealth interventions have the potential to help empower and engage traditionally marginalized populations in healthcare provision and research (23-30), then mHealth must be designed to serve these population’s needs, be accessible at the level of their understanding and abilities, and assist them in solving the problems they face during mHealth use (31, 32).

Several lessons emerged regarding the process of implementing a study using a digital health platform. First, we designed a rigorous pre-implementation plan to optimize the experience for participants and to boost full engagement from all team members. Specifically, each survey went through iterative rounds of review and testing. To promote readability, understanding, and brevity, the research team piloted each survey with community health workers, who were familiar with how survey questions could be best delivered to participants to collect the most informative responses. Feedback, particularly related to terminology and syntax, was then incorporated into the surveys. After translation of the survey into Spanish, this same process was followed. Second, the enrollment process was piloted at the PCC to ensure feasibility that study enrollment was not disruptive to clinician work-flow or a burden on patients. This process resulted in the integration of the CHW into morning huddles among clinicians, and established trust between stakeholders and confidence in the research team. Piloting at each step of implementation was crucial for rapid enrollment of participants and their sustained engagement with the study.

Still, there were logistical challenges during enrollment, including technology-related obstacles (e.g., owning a smartphone/tablet) and economic concerns (e.g., limited storage space, texting plans, and consistent access to WiFi or data plans to download the app and send/receive surveys). Not all potential participants approached owned a smart device or had access to email (e.g., many did not remember their email address and/or password, or did not know how to use email) to download the application from the store. In these cases, the CHW occasionally set up new email accounts and made wallet-sized cards with participants’ log-on information, but the time necessary for this may have resulted in some patients declining to participate in the study. To provide flexibility, the digital health platform used in this study was accessible via the web and as a mobile app. The platform also allowed for both email and text messaging options to receive surveys; therefore, participants were not necessarily required to have access to their email after enrollment (if using a smart device) or own a smart device (if using email) to access health data and to answer surveys. Further minimizing barriers to accessing the platform and surveys could be explored in the future, such as by utilizing alternative login methods (e.g., utilizing social media profiles, face ID, or fingerprint recognition), while recognizing that some of these alternative logins are associated with privacy concerns. Finally, the digital health app navigation system was in English and Spanish, though medical records connected from the patient portals were only in English – a limitation of current electronic health record systems,

Additionally, there were implementation challenges and real-life conceptual concerns that were revealed through the surveys. These related to having limited comfort with mHealth technology, mistrust of where health data are stored and who has access to that data, concerns about the security and privacy of that data, privacy policies, and historical perceptions of research participation. To understand and address these contextual factors, the CHW regularly reached out to participants to solicit any needs with navigating the digital health platform or completing surveys. The CHW also served as a resource for any questions or concerns regarding the privacy of the app. By maintaining a close relationship with the digital health company and research team through regular meetings, the CHW was able to communicate these questions and concerns to all stakeholders. This prompted updates to workflows and features of the platform in real-time that demonstrated a commitment to directly and promptly addressing participants’ concerns, thereby building trust. Moreover, the consistent interest indicated on all follow-up surveys in seeking the CHW for assistance, especially by the Spanish language-preferred participants, suggested that this population benefitted from additional attention and dialogue regarding research study participation and digital health platform use. It also reinforced that the CHW played a vital role in serving as a single approachable face of the study, as well as in listening and responding to concerns regarding the study and the platform.

Our study had several limitations. First, we only enrolled patients who had consistent access to a smart device, such as a smartphone or a tablet. Therefore, we do not know about the interest of using digital health platforms to monitor health and participate in research among those who do not own such devices. Second, we recruited English and Spanish language-preferred participants from two separate care facilities; thus, the differences observed between responses of the two groups may have been partially due to catchment area and the nature of the care facilities in addition to patient characteristics. Finally, we elicited qualitative feedback from a randomly-selected subset of patients, and these conclusions may not be applicable to the entire study sample or population at large.

## CONCLUSION

Despite several barriers, low-income individuals were feasibly enrolled in a mobile health app with the assistance of a CHW. Participants were interested in using various app features, including owning health records, monitoring health information, and participating in research.

Although they had concerns about safety and privacy, the involvement of a CHW facilitated engagement, trust, and participation through direct communication with all stakeholders. Designing mHealth apps and interventions must account for the unique needs and contexts of low-income, vulnerable populations to lessen health disparities and elevate the baseline health of these populations in new and innovative ways.

## Data Availability

All relevant data are within the paper and its Supplementary Materials files.

## ACKNOWLEDGEMENTS

We wish to thank the Hugo team for providing mobile health app and support, as well as the Aetna Foundation for funding the study.

